# Identification of a novel path for cerebrospinal fluid (CSF) drainage of the human brain

**DOI:** 10.1101/2023.01.13.23284147

**Authors:** Joel E. Pessa, Ronald M. Hoxworth

**Author notes:** (JEP).

## Abstract

How cerebrospinal fluid (CSF) drains from the human brain is of paramount importance to cerebral health and physiology. Obstructed CSF drainage may result in increased intracranial pressure and a predictable cascade of events including an enlarged subarachnoid space, dilated cerebral ventricles and ultimately cell death. The current and accepted model of CSF drainage in humans suggests CSF drains from the subarachnoid space into the sagittal sinus vein. Here we identify a new structure in the sagittal sinus of the human brain by anatomic cadaver dissection. The *CSF canalicular system* is the terminal CSF drainage of the human brain. It consists of paired channels on either side of the sagittal sinus vein that communicate directly with subarachnoid cerebrospinal fluid via virchow robin (perivascular) spaces. Fluorescent injection confirms that these channels are patent and that flow is independent of the venous system. Computer assisted tomography identified flow from the sagittal sinus to the cranial base. We verify our previous identification of CSF channels in the neck that travel from the cranial base to the subclavian vein. This information suggests a novel path for CSF drainage of the human brain and nerves that may represent the primary route for CSF recirculation. These findings have implications for basic anatomy, surgery, and neuroscience, and highlight the continued importance of gross anatomy to medical research and discovery.

## INTRODUCTION

How cerebrospinal fluid (CSF) drains from the human brain is of paramount importance to cerebral health and physiology. Obstructed CSF drainage may result in increased intra-cranial pressure and a predictable cascade of events including an enlarged subarachnoid space, dilated cerebral ventricles (hydrocephalus) and ultimately cell death (1, 2). This paper describes a new structure in the sagittal sinus of the human brain for CSF drainage, and confirms our previous findings of a CSF system in the neck. Together, this work suggests a novel path for CSF drainage of the brain that could represent the primary path for CSF recirculation in humans.

CSF is an ultra-filtrate of blood that has several functions (3–6). CSF produced in the ventricles serves as a reservoir across which the transport of energy molecules and solute occurs (3–6). CSF accumulates in the subarachnoid space where it acts as a shock absorber protecting the brain from impact.

The current and accepted model of CSF drainage in humans suggests CSF drains from the subarachnoid space into the sagittal sinus vein (Figs 1a and 1b) (7–10). CSF is thought to diffuse through protrusions of the arachnoid meninges called “arachnoid granulations” or “villi” (11). Although the number and distribution of arachnoid granulations is highly variable, this is the accepted route for CSF drainage in humans (12). CSF also drains to lymphatic vessels in the nose (via the cribiform plate) most likely as minor pathway. Researchers identified an alternative route in mice that involves CSF flow from dural lymphatics to the the scalp and neck (13, 14).

**Fig 1a.**
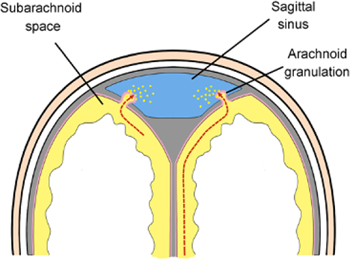
Left. Cross-section diagram of the accepted model of CSF drainage in humans. CSF (yellow) in the subarachnoid space is thought to diffuse through protrusions of the arachnoid meninges (arachnoid granulations) into the sagittal sinus vein (blue).

**Fig 1b.**
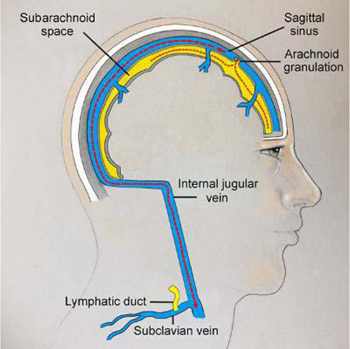
Right. Sagittal view of the current model of CSF drainage. CSF (yellow) drains from the subarachnoid space through arachnoid granulations into the sagittal sinus vein (blue). Sagittal sinus venous blood exits the cranium through the jugular foramen to enter the internal jugular vein.

Our previous work identified the CSF drainage of the neck (15). This present study identifies CSF channels in the sagittal sinus of the brain and completes the circuit for CSF drainage from the subarachnoid space to the subclavian vein.

## MATERIALS AND METHODS

Fresh cadaver dissections (not embalmed) were performed under institutional guidelines. The IRB committee reviewed this work and agreed that it does not require IRB approval or review. Seven (N = 7) dissections were performed in 5 male and 2 female cadavers. Ages ranged from 40 – 92 years (mean 82.5). The sagittal sinus was dissected to identify the bilateral CSF canalicular system, and photographs were obtained in selected specimens (Canon EOS 6D 50mm macro lens, Canon USA, Melville, NY). Computer assisted tomography (CT) was performed with the GE Helical CT Scan (GE Healthcare Systems, Chicago, Illinois) after the CSF canalicular system was intubated with 0.25 mm silastic tubing (WPI, Sarasota, Florida) and injected with 0.2 mls of Omnipaque^TM^ (iohexol). The CSF channels in the sagittal sinus were injected in 1 specimen using 1-5µ fluorescent polymer spheres (Cospheric, Santa Barbara, CA) after intubation with 22/24 gauge IV catheter (Baxter Healthcare Corporation, Deerfield, Illinois) and photographed under ambient/UV light. Neck dissections confirmed the anatomy of the CSF system in the neck in each specimen. One cervical biopsy was submitted for immunohistochemistry by 2-step indirect immunohistochemistry (IHC) for PROX-1, D2-40, CD 31, CD105, and F-actin (Sigma Aldrich, St. Louis, Missouri), and by direct IHC for the type-3 neurofilament protein vimentin (vimentin cy-3; Sigma Aldrich, St. Louis, Missouri) (15).

## RESULTS

Craniotomy exposed the dura and the midline venous sagittal sinus (Fig 2). After the sagittal sinus was opened, blood was evacuated to identify CSF channels located on either side of the venous sagittal sinus (Figs 3a and 3b).

**Fig 2.**
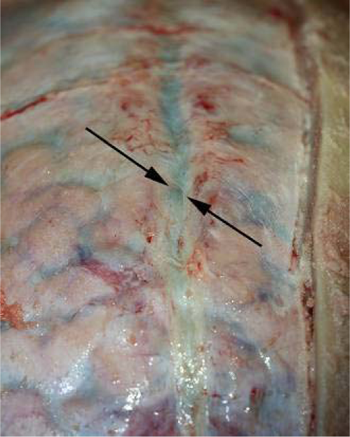
Demonstration of the venous sagittal sinus. The sagittal sinus is a midline venous structure (between arrows) in the dura. CSF channels travel on either side of this venous sinus.

**Fig 3a.**
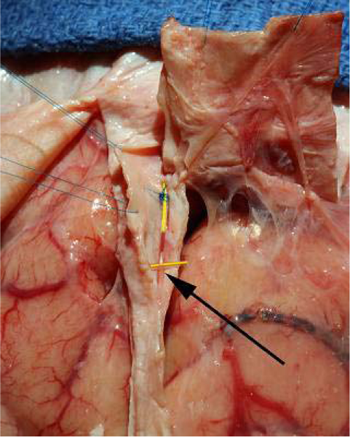
Left. CSF channels in the sagittal sinus. Blood was evacuated from the venous sagittal sinus and is held open with blue sutures. The right CSF channel system (arrow) is identified over the yellow marker in a 90’s year-old female.

**Fig 3b.**
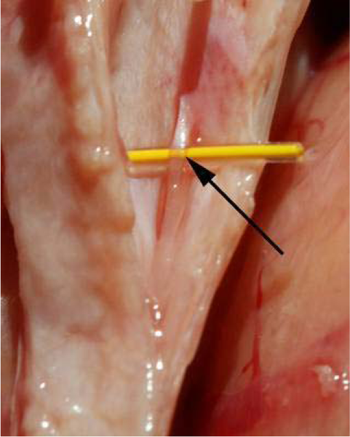
Right. CSF channels in the sagittal sinus. This is a macro view of CSF channels (arrow) in the sagittal sinus.

A cross-section view shows the location of CSF channels relative to the venous sinus (Fig 4a). CSF channels exist as a plexus that are located posteriorly and on either side of the venous system (Fig 4b). Dissection identified the CSF system in all specimens.

CT scan was performed in one specimen after the CSF channels on one side were injected with contrast material (Fig 5a). Helical CT scan confirmed flow in the CSF system (Fig 5b). Real-time imaging showed dye traveling from the superior sagittal sinus along the cranial base to exit the skull through the temporal fossa (Fig 5b). There was no extravasation of dye (Fig 5b), although CSF channels in the arachnoid meninges appeared to fill in a retrograde fashion (Fig 5b).

**Fig 4a.**
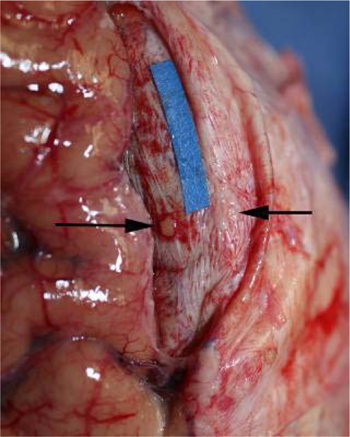
Left. Identification of the venous sagittal sinus. The sagittal sinus lies within the dura (beneath blue marker) of the falx cerebri. Tumor (black arrows) has metastasized to the dura and arachnoid meninges. 80’s year-old female.

**Fig 4b.**
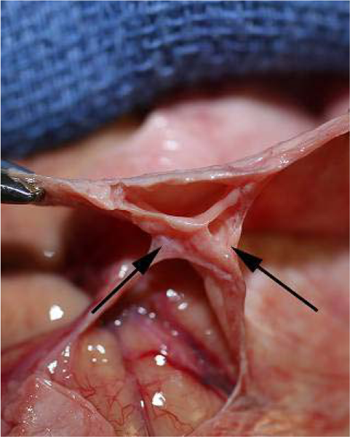
Right. CSF channels in the sagittal sinus. Clamps suspend dura and show the empty venous sinus.and bilateral CSF channels (arrows). The right side is widely patent, whereas the left has been obliterated by infiltrating carcinoma. The arachnoid meninges travel to these CSF channels.

**Fig 5a.**
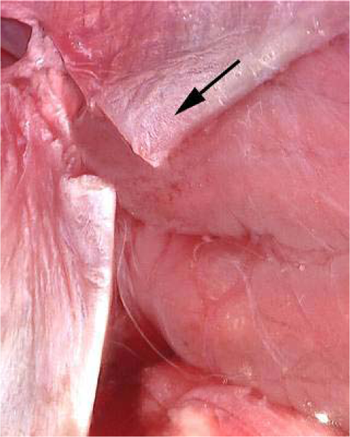
Left. Injection of CT contrast into CSF drainage. The CSF drainage was cannulated and injected with CT contrast (the direction of the injection was superior to inferior, arrow) in this 60’s year-old male specimen.

**Fig 5b.**
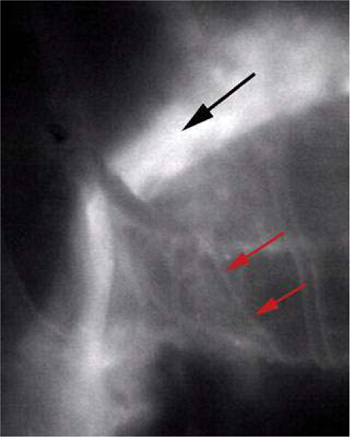
Right. CT imaging of CSF flow. CT scan shows contrast traveling in CSF channels from the sagittal sinus to the cranial base. There appears to be some retrograde flow into CSF channels in the arachnoid meninges (red arrows).

We used a fluorescent dye to confirm flow in one specimen. The CSF channels were identified and cannulated with intravenous catheters (Fig 6a) and Cospheric^TM^ dye was injected to show flow (Fig 6b). Imaging with UV light confirmed dye flow in the CSF system (Fig 6c). There appeared to be some back flow into the arachnoid meninges (Fig 6c). Dye injected into CSF channels did not fill the venous sagittal sinus or peripheral veins (Fig 6d).

**Fig 6a.**
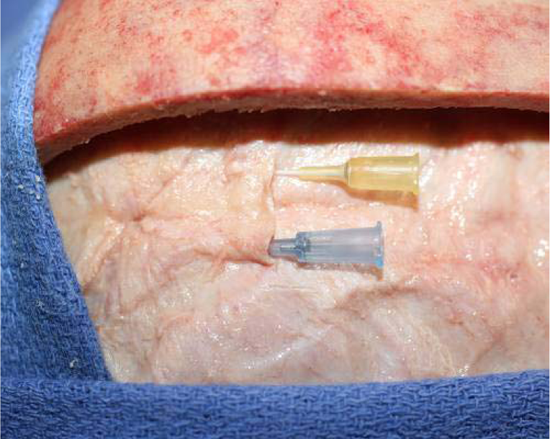
Left. Cannulation of CSF channels. The CSF channels were cannulated in this 60’s year old male specimen using a 22 gauge angiocatheter (blue) for the left system, and a 24 gauge angiocatheter (yellow) for the right system.

**Fig 6b.**
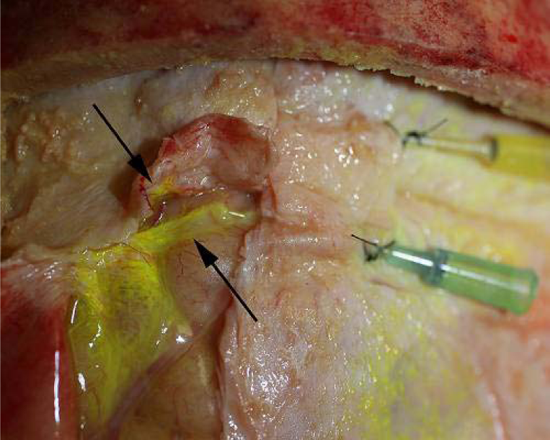
Right. Fluorescent dye injection. Cospheric^TM^ fluorescent polymer 1-5µ microparticles were injected into the CSF channels after which the sagittal sinus was transected. Flow was noted in both sides (arrows).

**Fig 6c.**
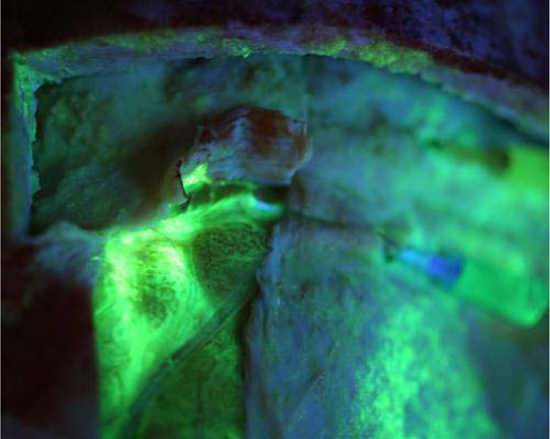
Left. Fluorescent imaging of flow in CSF channels. UV fluorescence shows documents flow within these CSF channels. Note the backflow into the arachnoid meninges (green fluorescent blush of tissue beneath left CSF system).

**Fig 6d.**
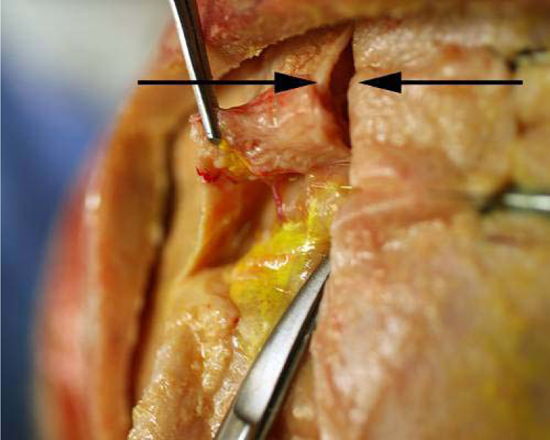
Right. CSF injection does not extravasate into veins. When the CSF channels were injected, there was no filling of the venous system. The venous sagittal sinus (black arrows) did not fluoresce. Note side branches (red vessels) of the sagittal sinus did not fill with fluorescent dye.

Immunohistochemistry was performed on a section of CSF channels harvested from the proximal neck. Sections were negative for lymphatic markers LYVE-1 (Fig 7a), and negative for vascular endothelial marker CD105 (Fig 7b). This specimen was positive for vimentin, and showed CSF channels travel as a plexus (Fig 7c).

**Fig 7a.**
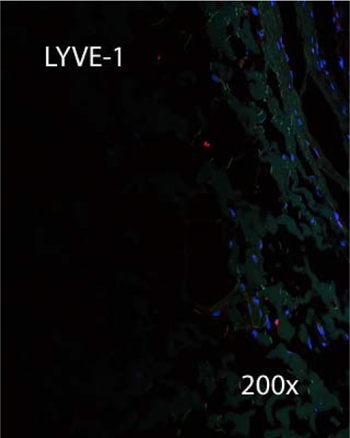
IHC for LYVE-1. IHC was negative for LYVE-1.

**Fig 7b.**
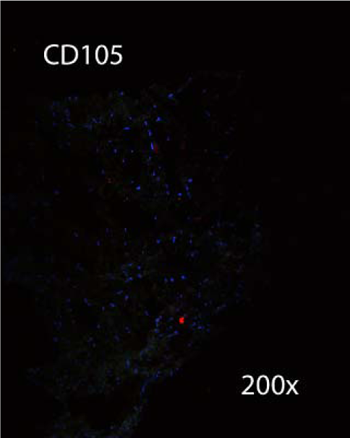
IHC for CD105. IHC was negative for vascular endothelial marker CD105.

**Fig 7c.**
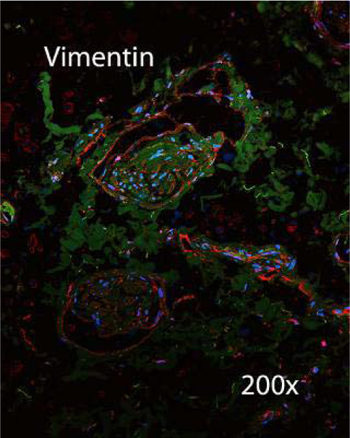
IHC for vimentin. IHC was positive for the type-3 neurofilament protein vimentin.

Anatomical dissection verified the previous finding of CSF channels in the neck. The cervical CSF system travels in the carotid sheath (Fig 8a). CSF channels are located in the outer layer (adventia) of the posterior internal jugular vein (Fig 8a). Another dissection identifies CSF channels as an additional structure traveling in the carotid sheath (Fig 8b).

**Fig 8a.**
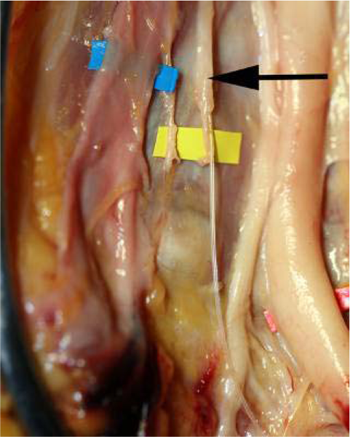
Left. Cervical CSF system. Neck dissection in 60’s year-old female specimen verifies the terminal CSF drainage in the neck (arrow). This travels within the adventia of the internal jugular vein (on blue marker). The carotid artery and vagus nerve are to the right of the CSF system.

**Fig 8b.**
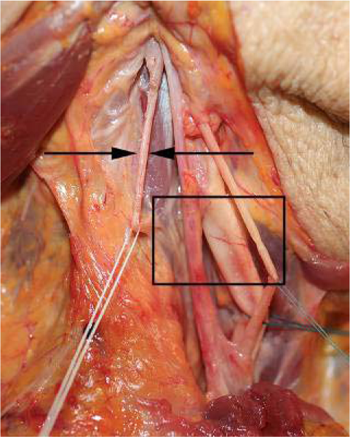
Right. Cervical CSF system. Neck dissection in an 80’s year-old female specimen verifies the terminal CSF drainage system in the neck (arrow). The structures of the carotid are seen in the rectangle and include (from left to right) the internal juglar vein, carotid artery, and vagus nerve.

## DISCUSSION

*The CSF canalicular system* is a novel anatomical path for CSF drainage of the human brain (Figs 9a and 9b). CSF channels in the superior sagittal sinus have not been previously described (7-12, 16-19). Because CSF channels travel as a group or plexus and are embedded in surrounding tissue, this system is named the CSF canalicular system. It is a consistent feature of sagittal sinus anatomy (Figs 10a and 10b).

**Fig 9a.**
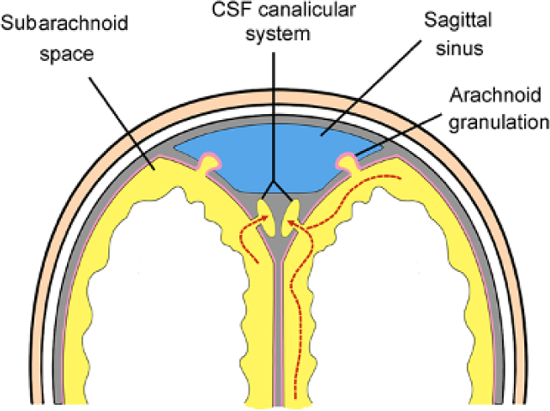
Left. A cross-section diagram of the CSF canalicular system. The CSF canalicular system is located on either side of the sagittal venous sinus. CSF (yellow) flows from the subarachnoid space into CSF channels.

**Fig 9b.**
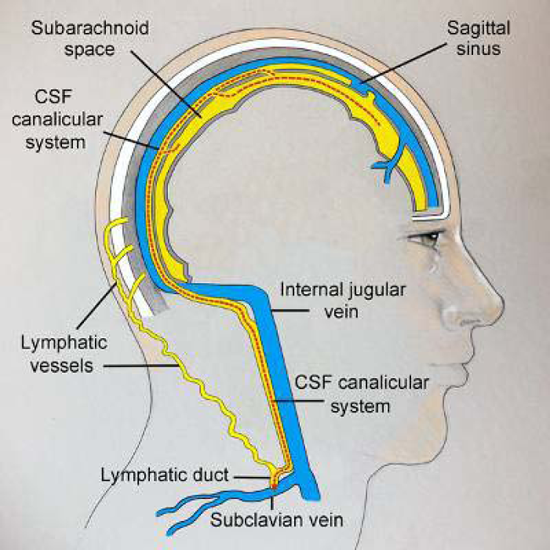
Right. Sagittal view of the CSF canalicular system. The CSF canalicular system provides an anatomical route for CSF (yellow) to drain from the subarachnoid space directly to the subclavian vein.

**Fig 10a.**
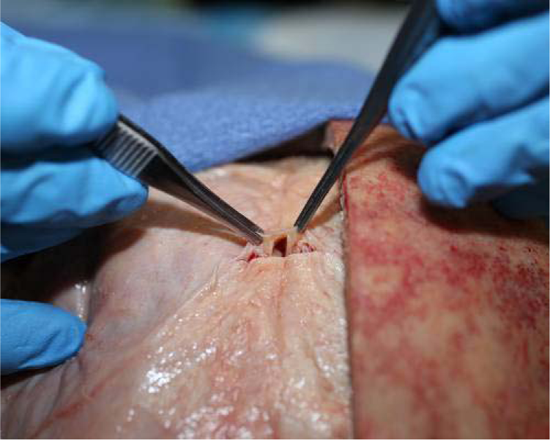
Left. The CSF canalicular system is a consistent feature of sagittal sinus anatomy. It is straightforward to identify the CSF canalicular system that is located on either side of the venous sagittal sinus.

**Fig 10b.**
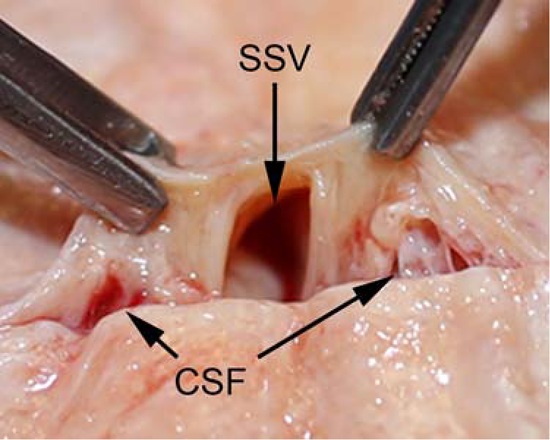
Right. The location of CSF canalicular system relative to the venous sagittal sinus. Macro view shows the centrally-located sagittal sinus vein (SSV) accompanied by CSF channels (CSF arrows) located on either side.

The CSF canalicular system is characterized as follows. CSF channels travel as a group or plexus, and appear as thin, translucent vessels that histologically lack a muscular layer. CSF drainage is privileged and does not involve intermediary lymphatic vessels or veins. CSF drains in a direct route from the arachnoid meninges to the subclavian vein, and has a similar terminal path to that described for peripheral nerves (15, 19). It is noteworthy that CSF channels in the neck were probably described by Cruickshank and Mascagni in 1786 and 1787 respectively, although CSF vessels were mischaracterized as lymphatics (20, 21). The features of the CSF canalicular system are summarized in Table 1.

**Table 1.**
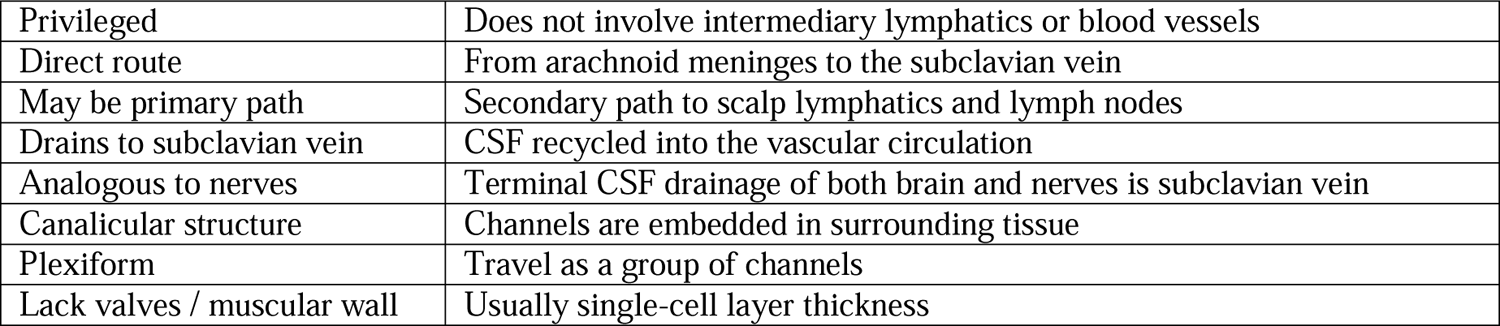
Characterization of the CSF canalicular system.

The limitation of this work is the same as any post-mortem anatomical study, and is unable to evaluate function. With that caveat, the authors hypothesize that the CSF canalicular system may be the primary path for CSF drainage in humans. Nerves are sensitive to pressure, and CSF drainage appears to be a redundant design in both the central and peripheral nervous systems (Figs 11a and 11b). The primary path for CSF drainage is humans may be via the CSF canalicular system, with a secondary path by extra-dural lymphatics to the scalp as described in mice (15, 22).

**Fig 11a.**
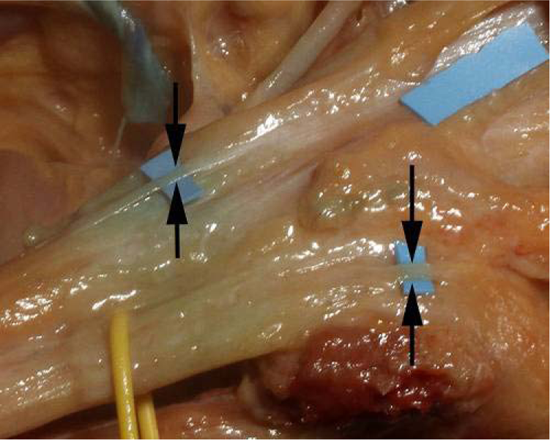
Left. The CSF canalicular system of brachial plexus. Draining CSF channels (arrows) on nerve (T1) travel within the outer layer of nerve (epineurium) that is embryologically analogous to the outer meninges of brain (dura). The terminal drainage of CSF in nerves is to the subclavian vein.

**Fig 11b.**
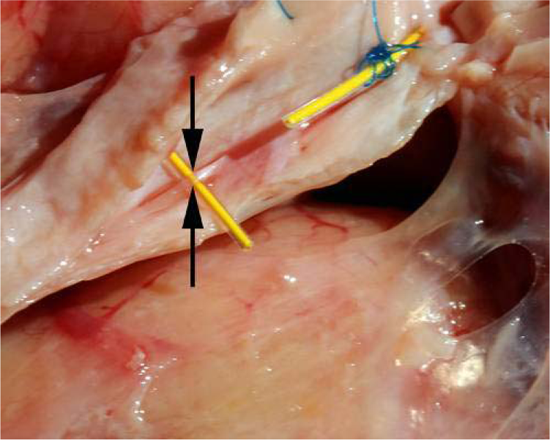
Right. Similarity of CSF channels in the central and peripheral nervous systems. Note the similarity in appearance of CSF channels in the sagittal sinus (arrow) to those found in human nerve (Fig 11a).

### CSF canalicular system is continuous with subarachnoid CSF via Virchow Robin spaces

Both our CT contrast injection (Fig 5b) and fluorescent injections (Figs 6b and 6c) suggest that the CSF canalicular system is continuous with subarachnoid CSF via Virchow Robin (VR) spaces. First identified in the 1850’s, these represent perivascular spaces through which interstitial fluid and CSF flow. A macro view of the fluorescent injection in particular shows (retrograde) filling of VR spaces located on either side of arachnoid veins (Figs 12a and 12 b). Immuohistochemistry suggests that glial cells may be associated with VR spaces in human arachnoid meninges (Fig 12c).

**Fig 12a.**
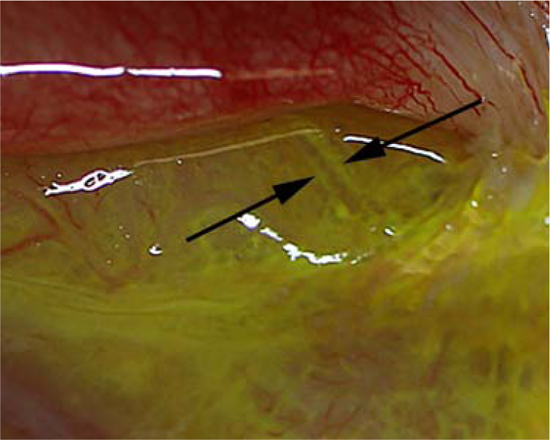
Left. Virchow Robin spaces in arachnoid meninges. Arrows show fluorescent dye in VR spaces on either side of an empty vein.

**Fig 12b.**
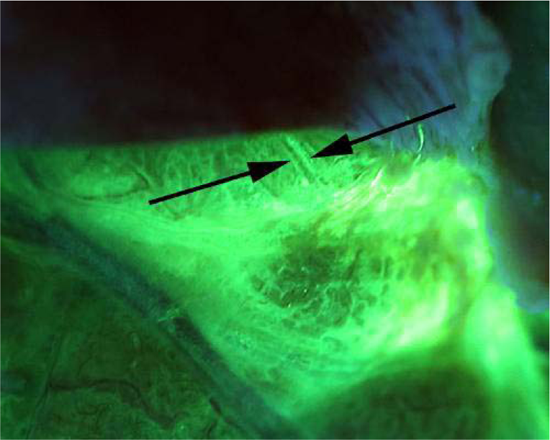
Right. Virchow Robin spaces. Fluorescence confirms dye entered VR spaces on either side of central vein after CSF canalicular injection.

**Fig 12c.**
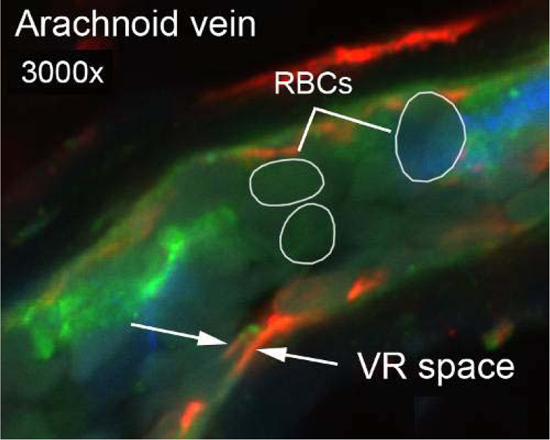
Glial cells may form VR spaces in human arachnoid meninges. IHC at 3000x of an arachnoid meningeal vein shows RBCs inside the vein lumen. Virchow Robin spaces surround the outer vein and are associated with glial cells (red fluorescence) in humans.

### The CSF canalicular system of brain is analogous to CSF drainage in nerves

The outer layer of nerve, epineurium, is embyrologically analogous to the dura of brain. In humans, CSF channels can be identified traveling in the epineurium (Fig 13). CSF channels in human peripheral nerves are morphologically similar to the CSF canalicular system (Figs 11a and 11b). The terminal CSF drainage of both brain and nerves is the subclavian vein. Like the brain, peripheral nerves appear to have redundant or secondary drainage routes to lymphatic vessels (Fig 13).

**Fig 13.**
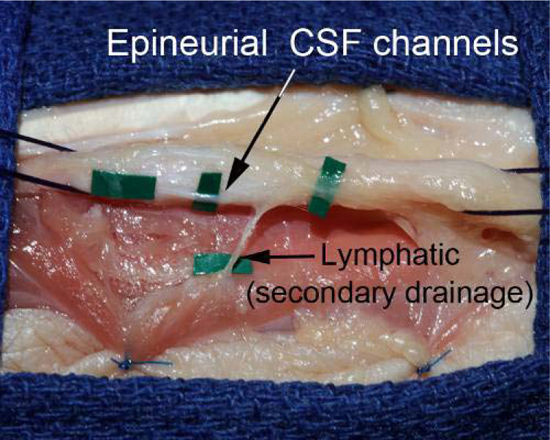
CSF canalicular system of human nerves. Epineurial CSF channels on human median nerve may be analogous to the CSF canalicular system of the brain. Primary drainage is to the subclavian vein; secondary drainage in case of obstruction is provided by CSF-lymphatic communications.

### Implications

The identification of the CSF canalicular system has implications for basic anatomy, surgery, and neuroscience. It redefines the anatomy of the sagittal sinus. The definition of what structures travel in the carotid sheath could be amended to include the terminal CSF drainage of the brain (23). Based on this study, Sappey’s Rule should be amended (24).

The “rule” that any vessel draining into a lymphatic (or into a lymph node) could be characterized as a lymphatic may not be accurate: lymphatics *and* CSF channels drain into the terminal lymphatic system (and then into the subclavian vein). This awaits further molecular characterization.

Evidence suggests that dysregulation of CSF drainage may be related to the etiology of neurocognitive disorders such as Alzheimer’s disease (25, 26). Certainly, dysregulation of flow in the CSF canalicular system may play a role in the etiology of communicating hydrocephalus (1, 2). The finding of subarachnoid enlargement would be predicted if there were acute obstruction of CSF outflow.

Astronauts subjected to prolonged microgravity may experience changes in vision, a disorder referred to as spaceflight-associated neuro-ocular syndrome (SANS). SANS is thought to be related to dysregulation of CSF flow (27–29). CSF drainage is gravity dependent. Cerebral ventricular volume is lower in the upright position (i.e. improved drainage) (30). The reverse is noted during spaceflight, where cerebral ventricular volume is increased in low-gravity environments (31). The increased thickness of the optic nerve sheath is explained by stasis of flow in CSF channels found in the optic nerve dura (32). Further research is needed (33–36).

## CONCLUSIONS

The CSF canalicular system is a novel path for cerebrospinal fluid drainage of the human brain. It is analogous to the path for CSF drainage on peripheral nerves. The identification of the CSF canalicular system has implications for anatomy, surgery, and neuroscience, and highlights the continued importance of gross anatomy to medical research and discovery.

## Data Availability

All data produced in the present study are available upon reasonable request to the authors.

## ACKNOWLEDGEMENTS

The authors thank the donors of the UTSW Willed Body Program and their families. Without their generous gift this research would not be possible. The authors thank the entire staff of the UTSW Willed Body Program for their help on this and all of our anatomical projects.

